# Large-scale screening of asymptomatic for SARS-CoV-2 variants of concern and rapid P.1 takeover, Curitiba, Brazil

**DOI:** 10.1101/2021.06.18.21258649

**Authors:** Douglas Adamoski, Jaqueline Carvalho de Oliveira, Ana Claudia Bonatto, Roseli Wassem, Meri Bordignon Nogueira, Sonia Mara Raboni, Edvaldo da Silva Trindade, Emanuel Maltempi Souza, SCB-UFPR COVID-19 team, Daniela Fiori Gradia

## Abstract

To provide a safer environment for individuals working on-site at the Federal University of Paraná, Curitiba, Brazil, we performed a large-scale mass testing SARS-CoV-2 program coupled with variant genotyping using two PCR-based approaches. We observed a fast dominance of the Gamma variant, displacing other variants in less than three months.

**Article Summary Line:** Coronavirus variants of concern may use asymptomatic population as silent spreaders to perform a fast displacement of previous established strains.

## Text

Continuous screening for SARS-CoV-2 active infections, coupled with contact tracing, could efficiently reduce the viral transmission among the community(1). However, as virus variants with increased transmission emerge, a rise of cases within affected regions is observed, demanding immediate actions for blocking virus spread and consistent and continuous genomic surveillance for continuously evaluating vaccination approches(2).

Brazil also showed consequences of the absence of genomic surveillance in a high seroprevalence scenario. Gamma (B.1.1.28/P.1) variant of concern (VOC) emerged in Manaus, Brazil, within three-quarters of the population positive for anti-N IgG, rapidly spreading over the city(3). This new variant was able to displace his predecessor, the B.1.1.28/P.2 variant of interest (VOI), previously the major lineage, among the wild one(4).

Extensive programs of routine asymptomatic screening for SARS-CoV-2 and follow-up sample genotyping are mandatory to control the cases and prevent further infection surges(5,6). The use of simplified approaches as multiplex qPCR provides a feasible, cost-effective way to discriminate samples and prioritize whole-genome sequencing efforts(5,7). In order to achieve this goal, we performed large-scale surveillance of infections and VOCs in the Federal University of Paraná community (students, technicians, professors, outsourced workers, cohabitants, and their relatives). The study was approved by the Research Ethics Committee (CAAE: 31687620.2.0000.0096).

## The study

In order to perform mass screening and genomic surveillance, from October 10th, 2020, to May 24th, 2021, asymptomatic and mild-symptomatic individuals within the community of Federal University of Paraná were called for voluntary participation by social media and e-mails. Saliva samples were collected in a pre-labeled 2.0 mL microtube using an individually wrapped plastic drinking straw, stored at 4°C storage until transport to the lab.

Each sample was homogenized and decanted for 30 minutes or centrifuged for 2 minutes (2.000 x g). Following this period, 200 µL from each specimen were pooled(8) in groups of five. RNA extraction was performed using an automated magnetic EXTRACTA - RNA and DNA Viral kit (Loccus Biotecnologia, Brazil). Amplification was performed on an QuantStudio5™ instrument (Thermo Fisher Scientific Inc., USA) using AllPlex nCov-2019 RT-PCR Master Mix Kit (SeeGene, South Korea) or Molecular SARS-CoV-2 EDx (Biomanguinhos/FioCruz, Brazil). The first kit include detection of the nucleocapsid (N), envelope (E), the RNA-dependent RNA polymerase (RdRP) virus genes, and a MS2-Phage spike-in as an internal control gene and the second detect E virus gene and human RNaseP. Some positive individuals were evaluated for antibodies against SARS-CoV-2 (IgM and IgG) after at least 15 days of RT-PCR result. Serum samples were performed by COVID-19 IgG / IgM ECO Test (Ecodiagnóstica Brasil) lateral flow immunochromatographic assay, according to the manufacturer’s instructions(9).

Positive samples were further evaluated using two probe-based genotyping systems to detect VOCs (Variants of Concern). The first one was the Vogels et al.(7) multiplex approach to detect Spike Δ69–70 and Orf1a Δ3675–3677 deletions as an outcome for distinguishing B.1.1.7, B.1.351 or P.1 and wild type or other lineages(7). To this approach, we also included de CDC N1 target and defined the threshold of Ct 28 to evaluate the gene dropouts. The second involved three allelic discrimination TaqMan assays (ThermoFisher, USA): N501Y (ANPRYZA), E484K (ANU7GMZ), and K417T (AN49ARF). The proposed readout was: P.1 (K417T, N501Y, and E484K), P.2 (only E484K), B.1.1.7 (only E484K), B.1.351 (N501Y and E484K, failure for K417T assay), and wild type/others for the absence of mutated alleles. This second assay had the discrimination power to distinguish the B1.1.28/P.2 from the wild type and the B.1.351/P.1. Both assays were performed using GoTaq® Probe 1-Step RT-qPCR System (Promega) on an QuantStudio5™ instrument (Thermo Fisher Scientific Inc., USA).

A total of sixteen collection dates were performed, with 12558 exams processed (Table 1), and 7249 people attended, since some participants engaged in more than one day of collection. The number of participants in each collection date varied due to more widespread dissemination within the community, increased service capacity, and engagement due to the perception of infection risk by the participants, ranging from 162 to 1737 attendees. The overall positivity rate was 1,28% (161/12558). Comparing those numbers to the Paraná state cases by epidemiological week of diagnosis (Figure A, blue bars), our positivity followed the beginning of state’s second SARS-CoV-2 infection wave (Figure A).

**Table 1.** Collection dates, engagement and positivity rates for SARS-CoV-2 infection.

| Date | Total | Positive (%) |
| --- | --- | --- |
| 02/10/2020 | 275 | 0 (0%) |
| 19/10/2020 | 279 | 0 (0%) |
| 06/11/2020 | 510 | 6 (1,18%) |
| 24/11/2020 | 1265 | 34 (2,69%) |
| 08/12/2020 | 1070 | 17 (1,59%) |
| 12/01/2021 | 1692 | 23 (1,36%) |
| 26/01/2021 | 1737 | 14 (0,81%) |
| 09/02/2021 | 1615 | 16 (0,99%) |
| 29/03/2021 | 196 | 1 (0,51%) |
| 12/04/2021 | 157 | 4 (2,55%) |
| 20/04/2021 | 872 | 2 (0,23%) |
| 26/04/2021 | 162 | 4 (2,47%) |
| 04/05/2021 | 884 | 12 (1,36%) |
| 10/05/2021 | 177 | 1 (0,56%) |
| 18/05/2021 | 1431 | 20 (1,4%) |
| 24/05/2021 | 236 | 7 (2,97%) |
| Total | 12558 | 161 (1,28%) |

**Figure.**
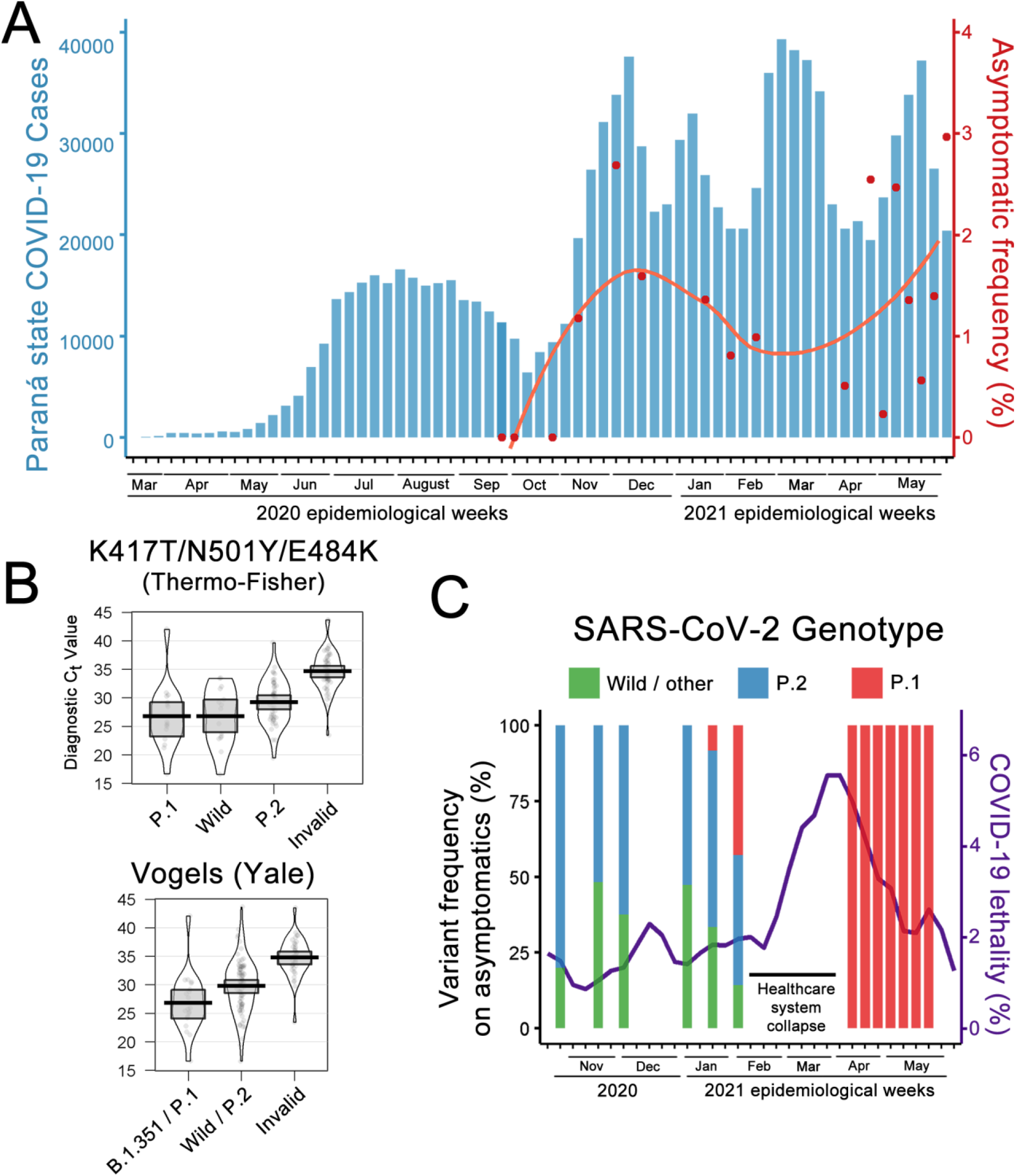
(A) Paraná state COVID-19 by diagnosis epidemiological week and UFPR’s mass testing program positive rates. Blue bars summarize positive cases in Paraná state accordingly to the diagnosis day, notified to state surveillance system until February, 15th 2021. Red dots represent the fraction of positive cases in all samples from mass screening collection date on UFPR, smoothed by LOESS (locally estimated scatterplot smoothing) in the red line. (B) Diagnostic Ct values and detection performance for Thermo-Fisher single plex (upper) and multiplex approach (lower). (C) COVID-19 Paraná state overall lethality (purple line) and variant prevalence among asymptomatic and mild-symptomatic cases by epidemiological weeks.

We performed a follow-up from 19 positive cases from December 4th (four participants) and December 18th (fifteen participants) with IgG and IgM serological tests. In only twice (one from each date), we observed negative results for IgG after 15 days from RT-PCR positive result; all cases returned negative for IgM. Seroconversion is not always observed after SARS-CoV-2 infection(10), raising questions concerning the test’s type and timing(11), besides the strength of the immune response in asymptomatic individuals(12). Both observed cases of seroconversion failure were detected with Ct values above 30 and one or two dropout targets in the multiplex RT-PCR (genes: E: non detected, RdRP: 34.7, N: 37.5 and genes: E: non detected, RdRP: non detected, N: 38.2), indicating a lower viral load in the test moment.

After February 2021, since the profile of the asymptomatic cases became unrelated to the state epidemiological curves, we started to evaluate retrospective and prospective all positive cases using multiplex and singleplex genotyping approaches. From all 161 positive cases were evaluated, Vogels et al. multiplex assay was invalidated in 46 (28.6%) against 50 (31.1%) in Thermo-Fisher three-assay allelic detection approach. Comparing the original Ct value of detection, there is a depreciation of performance in samples with Ct above 30 (Figure B), as stated in Thermo-Fisher manual. All cases were concordant between the two assays, considering that Vogels et al. alone do not discriminate between wild type and B1.1.28/P.2 VOI.

The first detection of the Gamma variant occurred on January 21th, 2021, less than two weeks after the Manaus city healthcare system collapsed. The Gamma variant appeared at 9.1% incidence on this date, increasing to 42.9% two weeks later (Figure C). This could be correlated to the following Curitiba city healthcare system collapse and a surge of COVID-19 lethality in the Paraná state, reaching values above 5% in the following weeks. When the testing activities returned, all cases became Gamma variant, completely displacing both B1.1.28/P.2 VOI and wild-type ones in three months.

## Conclusions

Analysis of saliva in pools in the present study presents a cheap, easy to collect, and feasible asymptomatic screening strategy. Thus, considering the transmission speed, until the vaccine is a large-scale option, the risk of asymptomatic COVID-19 spread should become a public awareness, focusing on social distancing. Our mass testing program was proposed to be accessible (since every test was free-of-charge for the participant), trusted (as every participant received its result and positive cases had a follow-up opportunity) and aimed to reach all social strata within the academic community (from professors to outsourced employees – mainly composed by social, economic and ethnic vulnerable groups), key characteristics to a strong mass testing system (13).

Both multiplex PCR and singleplex PCR approaches showed feasibility to evaluate the proportion of variants within genomic surveillance, with speed and lower cost than whole-genome sequencing (WGS) approaches. They do not replace WGS but could be an essential method to screen and select samples to further variant classification. Nevertheless, those approaches could show the fast-spreading of a new variant and predict COVID surges, acting as a lighthouse for wide-impact public health decisions.

## Data Availability

All data is available upon request

## Acknowledgments

We are grateful to Dr. Maria da Graça Bicalho for all the support, and a special thanks to the team of volunteers, since this job would not be possible without your help. This work was supported by the: PROPLAN /Federal University of Parana, Curitiba-Paraná-Brazil; FINEP - Funder of Studies and Projects, Ministry of Science, Technology and Innovation-Brazil - Institutional Network Project: Laboratories for Diagnostic tests for COVID-19 (0494/20).

## Author Bio

Dr. Adamoski is a substitute professor of genetics at the Curitiba Campus from Federal University of Paraná. His primary research interests include mRNA binding proteins, splicing regulation and genomics.

## SCB-UFPR COVID-19 team

Altina Bruna de Souza Barbosa; Beatriz Bocatte de Mattos; Bruna da Silva Soley; Carla Adriane Royer; Cibele Batina Rabelo; Cristina Kaehler; Diego Candido de Abreu; Guilherme Antonio Vendramin; Helyn Priscila de Oliveira Barddal; Letícia Dalla Vechia Henschel; Madson Silveira de Melo; Nathalie Carla Cardoso; Rachel dos Santos de Sena de Vasconcelos.

